# Risk factors for a low five-minute Apgar score at a provincial referral hospital in southern Mozambique: A secondary analysis of a matched case-control study

**DOI:** 10.64898/2026.10.13.26365388

**Authors:** Braiton Zefanias Maculuve, Natércia Fernandes

**Affiliations:** Faculty of Medicine, Eduardo Mondlane University, Maputo, Mozambique; National Directorate of Public Health, Ministry of Health, Maputo, Mozambique

## Abstract

Perinatal asphyxia is a leading cause of neonatal death in Mozambique and the main cause of early neonatal death in the referral hospitals of Gaza Province. Where blood gas analysis is unavailable, a low five-minute Apgar score remains a key marker of intrapartum compromise. This study aimed to identify maternal, obstetric, fetal and institutional factors associated with a low five-minute Apgar score at Xai-Xai Provincial Hospital, Mozambique. We conducted a secondary analysis of the records of 471 newborns delivered between January 2016 and December 2018, drawn from a matched case-control study of early neonatal death. The outcome was a five-minute Apgar score below 7. Associations were estimated with bivariate and multivariable conditional logistic regression and are reported as odds ratios with 95% confidence intervals. Of the 471 newborns, 76 (16.1%) had a low five-minute Apgar score. In the multivariable model, a gestational age of 42 weeks or more (adjusted odds ratio 4.04; 95% confidence interval 1.18–13.82) and ruptured membranes on admission (adjusted odds ratio 3.69; 95% confidence interval 1.37–9.96) were associated with higher odds of a low score, whereas partograph use during labour was associated with lower odds (adjusted odds ratio 0.07; 95% confidence interval 0.01–0.31). The association with preterm birth was attenuated after adjustment for birth weight. Post-term birth and ruptured membranes on admission were associated with a low five-minute Apgar score, and partograph use with lower odds. Because the sample was drawn from a study of early neonatal death, these associations should be confirmed in unselected birth cohorts. They nonetheless support strengthening intrapartum monitoring in Mozambican referral maternities.

## Introduction

In 2024, an estimated 4.9 million children died before their fifth birthday, 2.3 million of them in the neonatal period (the first 28 days of life), which represents 47% of all under-five deaths. Although the global under-five mortality rate fell to 37.4 deaths per 1,000 live births, less than half its level in 2000, progress has slowed markedly since 2015 [1].

If current trends continue, an estimated 28 million newborns will die between 2018 and 2030, most of them in sub-Saharan Africa and South Asia. Reducing neonatal mortality is essential to reduce under-five mortality, and the interventions needed to address the leading causes of neonatal death often differ from those needed for other deaths in children under five. Preventing intrapartum compromise, reflected in a low Apgar score, therefore contributes directly to the Sustainable Development Goal target for neonatal mortality [2].

The first minutes after birth are crucial for the newborn’s adaptation to extrauterine life. A low one-minute Apgar score has low sensitivity as a marker of asphyxia, whereas a low five-minute score is associated with increased mortality, particularly in preterm infants, and with later developmental and cognitive problems [3].

A low five-minute Apgar score indicates neonatal compromise and is associated with a higher risk of neonatal death. It also predicts adverse neurodevelopmental outcomes, including lower intelligence scores, motor impairment, attention-deficit/hyperactivity symptoms and autism [4–6].

The one-minute score reflects the condition of the newborn at birth and the response to resuscitation and has not been shown to be associated with long-term outcomes. In contrast, a low five-minute score confers an increased relative risk of cerebral palsy and childhood epilepsy, and a score below four is a stronger predictor of neonatal death than umbilical artery pH [7,8].

The diagnosis of perinatal asphyxia is a challenge in low-income countries because of the shortage of qualified human resources and of diagnostic tests. In these settings, the Apgar score is used to identify asphyxia; although the score alone is no longer accepted to define asphyxia, it remains an important indicator because it is easy to apply, and a score below seven at five minutes signals the need for special attention even in the absence of laboratory tests [9].

According to the annual report of the Mozambican national committee for the audit of maternal, perinatal and neonatal deaths, around 4,204 in-hospital neonatal deaths were recorded in 2019, an increase of 10% compared with 2018. The main causes were prematurity (39%), perinatal asphyxia (29%) and neonatal sepsis (15%). In Gaza Province, 115 neonatal deaths were recorded in the same period, and the main causes, in descending order, were perinatal asphyxia, prematurity and neonatal sepsis [10].

In many Mozambican health facilities, including those in Gaza Province, the diagnosis of perinatal asphyxia still relies mainly on the Apgar score. Despite the burden of asphyxia in the province, the factors associated with a low five-minute Apgar score have not been examined at Xai-Xai Provincial Hospital. This study aimed to identify maternal, obstetric, fetal and institutional factors associated with a low five-minute Apgar score in the maternity ward of Xai-Xai Provincial Hospital between 2016 and 2018.

## Materials and methods

### Study design and setting

This is a secondary analysis of data from a matched case-control study of maternal, fetal and institutional risk factors for early neonatal death, conducted in 2019 in five hospitals in Gaza Province, Mozambique: Xai-Xai Provincial Hospital (HPXX) and the rural hospitals of Chókwè, Chibuto, Chicumbane and Manjacaze. The primary study has not been published; its design is therefore described in full below. In the primary study dataset, 223 early neonatal deaths were recorded in these hospitals between 2016 and 2018; 142 (63.7%) occurred at HPXX, and perinatal asphyxia was the leading cause, followed by prematurity and neonatal sepsis.

The present analysis is restricted to newborns delivered at HPXX between 1 January 2016 and 31 December 2018. HPXX is the provincial referral hospital, located in Xai-Xai City, about 210 km north of Maputo; the city had approximately 141,963 inhabitants in the 2017 census. HPXX records approximately 4,000 deliveries per year, more than 90% of them from its primary catchment area. It was selected because it had the highest number of institutional deliveries in the province and the highest proportion of perinatal asphyxia (16%) among the five hospitals of the primary study.

### Participants

The primary study used a hospital-based case–control design and included births and early neonatal deaths recorded at the participating hospitals between 2016 and 2018. Cases were all early neonatal deaths (deaths within the first 7 days of life), identified through maternity ward registers, neonatal unit records and death registers.

Cases were excluded if they involved (i) congenital malformations considered incompatible with life; (ii) neonatal deaths referred from health areas outside the primary catchment area of the study health facilities; (iii) records classified by the study committee as invalid; (iv) deaths with a recalculated gestational age of less than 28 completed weeks; or (v) deaths following delivery outside the maternity facility.

Controls were selected from the same maternity ward registers and partographs used to identify eligible births. For each case, three surviving newborns delivered at the same hospital were selected using risk-set sampling. Controls were randomly selected among infants born on the same day as the case or within the 24-hour period immediately before or after the case birth. Controls were therefore matched to cases on hospital and date of birth. Newborns delivered outside the maternity facility were not eligible for selection as controls.

For neonatal deaths referred from other health facilities within the catchment area, additional controls were identified from the referring facilities to allow a sensitivity analysis of selection bias related to referral in the primary study. No cases were excluded because of an insufficient number of eligible controls.

Data on maternal, obstetric, fetal, and institutional characteristics were extracted from clinical records using a standardised data collection form completed by trained data collectors. The primary study included 223 cases and 669 controls (1:3 ratio); 142 cases and 336 controls had been delivered at HPXX.

Of the 478 newborns from HPXX included in the primary study, 7 had no recorded five-minute Apgar score and were excluded, leaving 471 newborns for the present analysis. Newborns transferred from health facilities outside the catchment area of the study hospitals were excluded; no newborns delivered outside the HPXX maternity ward or with congenital malformations were identified in the dataset. Because the sample size was determined by the available records, no formal sample size calculation was performed for this secondary analysis.

### Variables

The outcome was a low five-minute Apgar score, defined as a score below 7, as recorded in the delivery register by the attending health professional. Independent variables were grouped as maternal (age, occupation, parity, pre-eclampsia and eclampsia), obstetric (ruptured membranes on admission, fetal presentation, mode of delivery and prolonged labour), fetal (gestational age, birth weight, sex and twin pregnancy) and institutional (category of the professional who conducted the delivery and partograph use). Mode of delivery was classified as spontaneous vaginal or operative (caesarean section or instrumental vaginal delivery). Partograph use was defined as a partograph opened during labour. Ruptured membranes on admission were defined as rupture of the membranes already present on arrival at the maternity emergency room. Gestational age was classified as preterm (<37 weeks), term (37–41 weeks) or post-term (≥42 weeks), and birth weight as <2,500 g or ≥2,500 g. Missing values are reported for each variable in Table 1; observations with missing values for the variables included in a given model were excluded from that model (complete-case analysis).

**Table 1.** Maternal and obstetric characteristics by five-minute Apgar score, Xai-Xai Provincial Hospital, 2016–2018 (n = 471).

| Characteristic | Apgar $\geq 7$ (n = 395) | Apgar $< 7$ (n = 76) | Total (n = 471) |
| --- | --- | --- | --- |
| **Parity** |  |  |  |
| Nulliparous | 170 (43.0) | 35 (46.1) | 205 (43.5) |
| Multiparous | 225 (57.0) | 41 (53.9) | 266 (56.5) |
| **Pre-eclampsia** |  |  |  |
| No | 361 (91.4) | 62 (81.6) | 423 (89.8) |
| Yes | 32 (8.1) | 12 (15.8) | 44 (9.3) |
| Missing | 2 (0.5) | 2 (2.6) | 4 (0.8) |
| **Eclampsia** |  |  |  |
| No | 389 (98.5) | 69 (90.8) | 458 (97.2) |
| Yes | 2 (0.5) | 5 (6.6) | 7 (1.5) |
| Missing | 4 (1.0) | 2 (2.6) | 6 (1.3) |
| **Ruptured membranes on admission** |  |  |  |
| No | 318 (80.5) | 48 (63.2) | 366 (77.7) |
| Yes | 66 (16.7) | 23 (30.3) | 89 (18.9) |
| Missing | 11 (2.8) | 5 (6.6) | 16 (3.4) |
| **Fetal presentation** |  |  |  |
| Cephalic | 377 (95.4) | 65 (85.5) | 442 (93.8) |
| Breech | 5 (1.3) | 2 (2.6) | 7 (1.5) |
| Missing | 13 (3.3) | 9 (11.8) | 22 (4.7) |
| **Mode of delivery** |  |  |  |
| Spontaneous vaginal | 342 (86.6) | 61 (80.3) | 403 (85.6) |
| Operative (caesarean or instrumental) | 53 (13.4) | 15 (19.7) | 68 (14.4) |
| **Prolonged labour** |  |  |  |
| No | 386 (97.7) | 63 (82.9) | 449 (95.3) |
| Yes | 6 (1.5) | 12 (15.8) | 18 (3.8) |
| Missing | 3 (0.8) | 1 (1.3) | 4 (0.8) |

Values are n (%); column percentages calculated over all newborns in each group, including missing values.

### Statistical analysis

Data were analysed with R version 4.0.3 [11]. The distribution of each variable, implausible values and missing data were first examined graphically. Categorical variables are presented as absolute and relative frequencies by outcome group. Because participants were sampled in matched sets, associations were estimated with conditional logistic regression. Each independent variable was first examined in a bivariate model.

Variables were then examined in separate multivariable models for each group of factors (newborn, obstetric and institutional characteristics). Because of the limited number of outcome events, the final multivariable model included a limited set of variables representing these groups: newborn sex and birth weight, gestational age, ruptured membranes on admission, twin pregnancy and partograph use. The category of the professional who conducted the delivery was not included because of probable confounding by indication. No subgroup, interaction or sensitivity analyses were performed. Results are reported as crude and adjusted odds ratios (OR) with 95% confidence intervals (CI); a two-sided p ≤ 0.05 was considered statistically significant. Full model results are provided in S1 Table. The study is reported following the STROBE guidelines [12] (S1 Checklist).

### Ethics statement

The primary study was approved by the National Bioethics Committee for Health of Mozambique (CNBS; registration 87/CNBS/18, reference 399/CNBS/18). The protocol of the present analysis was approved by the Institutional Bioethics Committee for Health of the Faculty of Medicine and Maputo Central Hospital (CIBS FM&HCM; reference 097/2020). Because this was a retrospective analysis of anonymised secondary data routinely collected at the HPXX maternity ward, individual informed consent was not obtained, as approved by the committee. The authors did not have access to information that could identify individual participants. Authorisation to use the dataset was obtained from the investigators of the primary study, and a cover letter was issued by the Gaza Provincial Health Directorate. The study was conducted in accordance with the Declaration of Helsinki.

## Results

Of the 471 newborns, 76 (16.1%) had a low five-minute Apgar score. Mean maternal age was 24.7 years (SD 6.3). Of the 470 mothers with known age, 343 (73.0%) were aged 19–34 years, 84 (17.9%) were younger than 19 years and 43 (9.1%) were 35 years or older. Of the 425 women with information on occupation, 366 (86.1%) were not in paid employment.

Overall, 205 women (43.5%) were nulliparous and 98 of 467 (21.0%) were classified as being at high obstetric risk; 44 of 467 (9.4%) had pre-eclampsia and 7 of 465 (1.5%) eclampsia. Membranes were ruptured on admission in 89 of 455 women (19.6%) (Table 1). Among the newborns with known gestational age, 87 of 462 (18.8%) were preterm and 43 (9.3%) were post-term; 22 of 471 (4.7%) were from twin pregnancies and 90 of 468 (19.2%) weighed less than 2,500 g.

In bivariate analysis (Table 2), preterm and post-term birth, pre-eclampsia, ruptured membranes on admission and delivery by a higher-level professional were associated with higher odds of a low five-minute Apgar score, whereas a birth weight of 2,500 g or more, maternal paid employment and partograph use were associated with lower odds.

**Table 2.** Bivariate conditional logistic regression of factors associated with a low five-minute Apgar score, Xai-Xai Provincial Hospital, 2016–2018.

| Variable | Crude OR | 95% CI | p-value |
| --- | --- | --- | --- |
| **Gestational age (ref. 37–41 weeks)** |  |  |  |
| <37 weeks | 10.33 | 4.06–26.30 | <0.001 |
| ≥42 weeks | 3.07 | 1.13–8.40 | 0.029 |
| **Birth weight (ref. <2,500 g)** |  |  |  |
| ≥2,500 g | 0.04 | 0.01–0.14 | <0.001 |
| **Maternal age (ref. 19–34 years)** |  |  |  |
| <19 years | 1.17 | 0.56–2.46 | 0.675 |
| ≥35 years | 0.54 | 0.17–1.72 | 0.295 |
| **Mother in paid employment (ref. no)** |  |  |  |
| Yes | 0.25 | 0.07–0.87 | 0.029 |
| **Pre-eclampsia (ref. no)** |  |  |  |
| Yes | 2.72 | 1.13–6.51 | 0.025 |
| **Mode of delivery (ref. spontaneous vaginal)** |  |  |  |
| Operative (caesarean or instrumental) | 2.39 | 0.94–6.05 | 0.067 |
| **Ruptured membranes on admission (ref. no)** |  |  |  |
| Yes | 2.24 | 1.13–4.46 | 0.022 |
| **Twin pregnancy (ref. singleton)** |  |  |  |
| Yes | 4.23 | 0.99–18.04 | 0.051 |
| **Professional who conducted delivery (ref. mid- or basic-level)** |  |  |  |
| Obstetrician-gynaecologist or higher-level technician | 14.78 | 3.30–66.20 | <0.001 |
| **Partograph use (ref. no)** |  |  |  |
| Yes | 0.06 | 0.02–0.17 | <0.001 |

OR, odds ratio; CI, confidence interval; ref., reference category. Crude odds ratios from conditional logistic regression accounting for the matched design.

Compared with term newborns, preterm newborns had ten times higher odds of a low score (OR 10.33; 95% CI 4.06–26.30) and post-term newborns three times higher odds (OR 3.07; 95% CI 1.13–8.40). Newborns weighing 2,500 g or more had lower odds than those weighing less (OR 0.04; 95% CI 0.01–0.14). Twin pregnancy was associated with higher odds of a low score, although the confidence interval included 1 (OR 4.23; 95% CI 0.99–18.04). Maternal age was not associated with the outcome: compared with mothers aged 19–34 years, the OR was 1.17 (95% CI 0.56–2.46) for mothers younger than 19 years and 0.54 (95% CI 0.17–1.72) for those aged 35 years or older.

Newborns of mothers in paid employment had lower odds of a low score (OR 0.25; 95% CI 0.07– 0.87). Operative delivery (caesarean section or instrumental vaginal delivery) was not significantly associated with a low score (OR 2.39; 95% CI 0.94–6.05), whereas maternal pre-eclampsia was (OR 2.72; 95% CI 1.13–6.51). Deliveries conducted by an obstetrician-gynaecologist or a higher-level health technician were associated with markedly higher odds of a low score than those conducted by mid- or basic-level staff (OR 14.78; 95% CI 3.30–66.20). Partograph use was associated with lower odds of a low score (OR 0.06; 95% CI 0.02–0.17).

The multivariable model included 424 newborns with complete data, of whom 65 had a low five-minute Apgar score (Table 3). Post-term birth remained associated with a low score (adjusted OR 4.04; 95% CI 1.18–13.82), whereas the association with preterm birth was attenuated and no longer statistically significant (adjusted OR 2.18; 95% CI 0.39–12.09). Birth weight of 2,500 g or more (adjusted OR 0.17; 95% CI 0.03–1.04) and twin pregnancy (adjusted OR 1.53; 95% CI 0.15– 16.14) were not significantly associated with the outcome. Ruptured membranes on admission were associated with higher odds of a low score (adjusted OR 3.69; 95% CI 1.37–9.96), and partograph use remained associated with lower odds (adjusted OR 0.07; 95% CI 0.01–0.31).

**Table 3.** Multivariable conditional logistic regression of factors associated with a low five-minute Apgar score, Xai-Xai Provincial Hospital, 2016–2018.

| Variable | Adjusted OR | 95% CI | p-value |
| --- | --- | --- | --- |
| **Gestational age (ref. 37–41 weeks)** |  |  |  |
| <37 weeks | 2.18 | 0.39–12.09 | 0.373 |
| ≥42 weeks | 4.04 | 1.18–13.82 | 0.026 |
| **Birth weight (ref. <2,500 g)** |  |  |  |
| ≥2,500 g | 0.17 | 0.03–1.04 | 0.055 |
| **Ruptured membranes on admission (ref. no)** |  |  |  |
| Yes | 3.69 | 1.37–9.96 | 0.010 |
| **Twin pregnancy (ref. singleton)** |  |  |  |
| Yes | 1.53 | 0.15–16.14 | 0.723 |
| **Partograph use (ref. no)** |  |  |  |
| Yes | 0.07 | 0.01–0.31 | <0.001 |

OR, odds ratio; CI, confidence interval; ref., reference category. Adjusted odds ratios from a multivariable conditional logistic regression model including all variables shown and newborn sex (n = 424; 65 events).

## Discussion

In this secondary analysis, 16.1% of newborns had a low five-minute Apgar score. After adjustment, post-term birth and ruptured membranes on admission were associated with higher odds of a low score, and partograph use with lower odds.

Post-term newborns had four times higher adjusted odds of a low score. Placental insufficiency and cord compression secondary to oligohydramnios are plausible mechanisms [13], and a similar association was reported in two Swedish hospitals [14]. In a matched case-control study in Cali, Colombia, induction of labour was associated with markedly lower odds of perinatal asphyxia [15], which suggests that timely induction in post-term pregnancies may be protective, although its implementation depends on the capacity of each facility.

Ruptured membranes on admission were associated with more than three times higher odds of a low score. Rupture of membranes increases the risk of chorioamnionitis, neonatal sepsis, abnormal presentation and placental abruption, and prolonged rupture has been linked to intraventricular haemorrhage and adverse neurodevelopment. The duration of rupture and signs of infection were not available in the analysis dataset, which limits interpretation. A similar association was reported in a low-income population in the United States [16].

Partograph use was strongly associated with lower odds of a low score. The partograph, promoted by the World Health Organization as an early-warning tool for abnormal progress of labour [17], may support timely decisions on referral and delivery. However, in routine records a completed partograph may also reflect better overall quality of intrapartum care or more complete documentation, and women admitted in advanced labour or with complications may be less likely to have one opened. The magnitude of this association should therefore be interpreted with caution. Preterm birth showed the strongest crude association, but this was attenuated after adjustment for birth weight, which is consistent with the close correlation between the two variables. Prematurity and low birth weight are consistently reported as determinants of a low five-minute Apgar score in Ethiopian hospitals [2,3,18,19], and pre-eclampsia has been associated with low scores elsewhere [20]. Because the five-minute score is a strong predictor of neonatal mortality [21], these factors remain relevant targets for intrapartum and neonatal care.

The higher odds of a low score among deliveries conducted by obstetricians or higher-level technicians most likely reflect confounding by indication, since complicated labours are referred to the most qualified staff. This variable was therefore not interpreted as a risk factor and was not included in the multivariable model.

## Strengths and limitations

This study used data from the main referral maternity of Gaza Province, where perinatal asphyxia is the leading cause of early neonatal death. Several limitations should be considered. First, participants were drawn from a matched case-control study of early neonatal death; because neonatal death is strongly associated with a low Apgar score, the sample over-represents compromised newborns, and the observed proportion does not reflect the hospital population. Second, the data were collected retrospectively from clinical records, with missing information for some variables, and the dataset did not include umbilical cord pH, lactate, duration of membrane rupture, fetal heart rate monitoring, maternal education or macrosomia. Third, Apgar scores were assigned by different professionals without standardisation. Fourth, the small number of events produced wide confidence intervals for some estimates, such as twin pregnancy. Finally, the data refer to 2016–2018 and to a single hospital, which limits generalisability.

## Conclusions

Post-term birth and ruptured membranes on admission were associated with a low five-minute Apgar score at Xai-Xai Provincial Hospital, whereas partograph use was associated with lower odds. These findings support the timely management of post-term pregnancies, consistent partograph use and early assessment of women with ruptured membranes in Mozambican referral maternities. Prospective studies in unselected birth cohorts are needed to confirm these associations.

## Data Availability

All relevant data supporting the findings of this study are within the manuscript and its Supporting Information files.

## Acknowledgments

The authors thank Dr Mulássua José Simango, principal investigator of the primary study, for authorising the use of the database, and Dr Arianna Rosales, Dr Belkis Machado and Dr Sandra Gaveta for their methodological guidance.

## Supporting information

**S1 Checklist. STROBE checklist for case-control studies**. Completed checklist with the page and line numbers of each reported item.

**S1 Table. Full results of the bivariate and multivariable conditional logistic regression models**. Regression coefficients, standard errors, odds ratios, 95% confidence intervals, p-values and goodness-of-fit measures.

